# Genetic Evaluation of the Role of Galactose-Deficient IgA1 in IgA Nephropathy: Evidence from Association Testing and Mendelian Randomization

**DOI:** 10.1101/2025.10.25.25338806

**Authors:** Winston W. S. Fung, Gabriel T. Doctor, Omid Sadeghi-Alavijeh, Jingyi Wu, Sigrid Lundberg, Jicheng Lv, Hong Zhang, Xu-Jie Zhou, Xueqing Yu, Jonathan Barratt, Daniel P. Gale

**Affiliations:** Department of Renal Medicine, University College London, London, UK; Renal Division, Peking University First Hospital, Peking University Institute of Nephrology, Beijing, China; Department of Medical Specialist Care, Nephrology Clinic and Department of Clinical Sciences, Karolinska Institutet, Danderyd University Hospital, Stockholm, Sweden; Department of Nephrology, Guangdong Provincial People’s Hospital, Guangzhou, Guangdong, China; Department of Cardiovascular Sciences, University of Leicester, Leicester, UK

## Abstract

Elevated galactose deficient IgA1 (Gd-IgA1) is known to be associated with IgA nephropathy (IgAN) and is often regarded as the first of four steps in the “four hit” hypothesis to explain disease pathogenesis. However, while the proposed downstream hits have support from unbiased genetic association studies, similar genetic evidence to support a causal role for Gd-IgA1 in the disease is lacking. Multiple previous genome-wide association studies have shown that common variation in the gene *C1GALT1* is strongly associated with Gd-IgA1 levels. We used this established relationship first to calculate power to detect an association of *C1GALT1* alleles with IgAN risk and second to conduct association and Mendelian randomisation analyses to detect and quantify evidence for a causal role of Gd-IgA1 in IgA nephropathy. Despite adequate power, we did not observe significant genetic evidence for a causal role of Gd-IgA1 in IgAN that would explain the well-established observational association, and infer that this may not be explained by causation. This raises the possibility that Gd-IgA1 might better be regarded as a biomarker than a cause of IgA nephropathy and suggests that caution is needed when inferring clinical efficacy of treatments based on their effects on Gd-IgA1 levels.

**Lay Summary:** Immunoglobulin A (IgA) nephropathy is a kidney disease in which IgA builds up in the kidneys, causing inflammation there. People with the condition often have higher levels of a form of IgA that lacks a sugar called galactose (galactose-deficient IgA1, or Gd-IgA1) but it has been unclear whether this abnormal IgA actually causes the disease, or whether it is simply a marker of it. To address this, we used two genetic experiments: first, we tested whether people who inherit genetic variants that increase Gd-IgA1 levels are more likely to develop IgA nephropathy; and second, we used Mendelian randomization methods that compare the genetic effects on Gd-IgA1 levels with their effects on disease risk. Despite adequate power, neither approach showed that Gd-IgA1 causes IgA nephropathy, suggesting the raised levels seen in patients might be a consequence of other processes. This means treatments should not be judged solely on whether they lower Gd-IgA1.

## Introduction

IgA nephropathy (IgAN) is the most prevalent primary glomerulonephritis worldwide with a high lifetime risk of kidney failure [1]. The “four-hit hypothesis” has been developed to explain the pathogenesis of IgAN [2]. The proposed “first hit” is the over-production of galactose-deficient IgA1 (Gd-IgA1) leading to elevated circulating levels. This is suspected to trigger Gd-IgA1 specific autoantibody production (“second hit”) with immune-complex formation (“third hit”) and downstream deposition in the kidney leading to glomerular inflammation (“fourth hit”) [2]. However, while genome wide association studies (GWAS) have provided evidence for hits two to four [3], genetic evidence supporting a causal role of Gd-IgA1 overproduction in IgAN has remained elusive.

Human IgA1 undergoes *O*-linked glycosylation at serines and threonines within its hinge region. These glycans comprise combinations of *N*-acetylgalactosamine, galactose and sialic acid moieties. Chains lacking galactose (termed galactose-deficient IgA1, Gd-IgA1) are more abundant in the serum of patients with IgAN than in controls. Longitudinal and family-based analyses of Gd-IgA1 levels in humans have shown that Gd-IgA1 level is highly heritable (h^2^=0.387) and stable over time in individuals [4]. Across multiple geographical regions, higher Gd-IgA1 levels have been observed in patients with IgAN compared with controls. In observational case-control studies, each standard deviation (SD) increase in Gd-IgA1 is associated with an odds ratio (OR) of IgAN of approximately 1.5 (95% Confidence Intervals (CIs) all between 1.31 and 1.78) [4,5]. In addition, in a randomised, placebo-controlled clinical trial of Nefecon therapy in IgAN, reduction in serum Gd-IgA1 levels alongside reductions in both IgA1 and IgA/IgG immune complex levels were reported in those receiving the drug compared with placebo [6]. These data have supported the hypothesis that elevated Gd-IgA1 levels may play a causal role in the development of IgAN.

Paradoxically, although higher circulating Gd-IgA1 levels are consistently associated with IgAN, mean levels are often higher in European populations than in East Asian populations, even when measured side-by-side in the same laboratory [4], despite IgAN being more prevalent in the latter—raising the possibility that the strong association with IgAN may not result from Gd-IgA1 playing a causal role in the disease and could instead be explained by factors that differ by population or geography, are confounded by other variables, or even by reverse causation.

Genome wide association studies (GWAS) of Gd-IgA1 levels have revealed strong and statistically robust signals at *C1GALT1*, *C1GAT1C1* and *GALTNT12* (which together encode key proteins necessary for IgA1 galactosylation) in individuals with IgAN and in healthy subjects, with the set of alleles at these loci associated with elevated Gd-IgA1 being more common, and explaining more of the observed variation, in European compared with East Asian cohorts (Table S1) [4,5,7]. Although the total amount of variability in Gd-IgA1 levels explained by common variation at *C1GALT1* is small (4% and 1-4% of the variance in Europeans and Chinese cohorts respectively [4,5,7]) these studies provided insight into mechanisms contributing to the heritability of this trait, and quantification of the genetic associations can be used to address the question of the extent to which elevated Gd-IgA1 plays a causal role in the pathogenesis of IgAN: if Gd-IgA1 causes IgAN, genetic risk factors for elevated Gd-IgA1 would necessarily also make a contribution to the risk of IgAN.

In the largest and most recently reported GWAS in IgAN [3], no evidence of association with IgAN was detected at *C1GALT1* in European or East Asian cohorts analysed separately or in a biethnic metanalysis (Figure 1). We sought to address the question of whether this lack of association is explained by inadequate power to detect an association. We genotyped additional samples at the *C1GALT1* to obtain additional power and analysed the relationship using a Mendelian Randomisation framework, adequately powered to detect a causal relationship between *C1GALT1* levels and IgAN, to give a robust effect size estimate.

**Figure 1.**
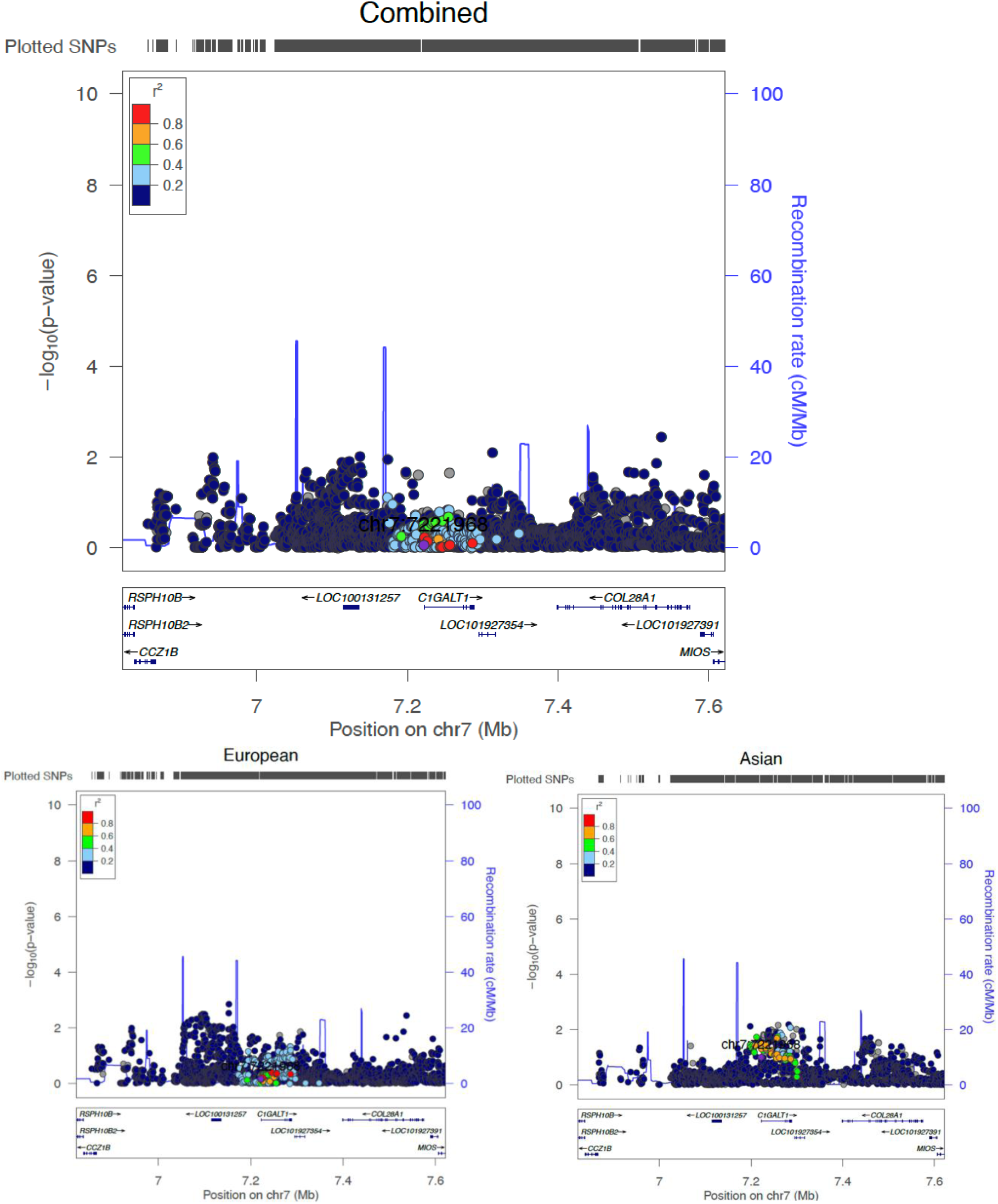
Association of SNPs across *C1GALT1* with IgA nephropathy [3] is shown, with Gd-IgA1-associated SNP rs758263 highlighted in purple and markers in LD with this variant coloured as shown.

## Methods

### Data Sources

We accessed the summary statistics from the largest recently published biethnic IgAN GWAS (10,146 cases and 28,751 controls) [3]. We genotyped 10,079 Chinese individuals with IgAN together with 5,030 geographically matched unrelated healthy subjects at 14 SNPs across the *C1GALT1* locus selected to capture the known Gd-IgA1 associated alleles in European and Chinese populations using KASP assays. Together with the biethnic IgAN GWAS, the combined sample size was 54,006 individuals, including 27,272 Chinese individuals. Summary statistics of GWAS of Gd-IgA1 levels were obtained from a Caucasian cohort (513 samples) [4] and a Chinese cohort (1162 samples) [7]. We focused our genotyping on the *C1GALT1* locus and not the X chromosomal *C1GALT1C1* given the greater availability of the summary statistics for associations at the *C1GALT1* (autosomal) locus, and the far stronger effect of variation at *C1GALT1* on Gd-IgA1 levels means that it is a more powerful genetic instrument (Table S1). Table 1 summarises the datasets included in this study.

**Table 1.** Exposure (Gd-IgA1) and Outcome (IgAN) cohorts included in this study

| Cohort | Number | Case proportion | Source of data |
| --- | --- | --- | --- |
| Gd-IgA1 studies (MR Exposure) |  |  |  |
| British_2017 | 513 | 1.00 | [4] |
| European_2017 | 1030 | 1.00 | [5] |
| East_Asian_2017 | 1603 | 1.00 | [5] |
| Chinese_2021 | 1162 | 1.00 | [7] |
| IgAN studies (MR Outcome) |  |  |  |
| European_2023 | 26734 | 0.42 | [3] |
| East_Asian_2023 | 12163 | 0.38 | [3] |
| Chinese_2025 | 15109 | 0.67 | not previously published |

### IgAN GWAS Meta-analysis

Testing for association with IgAN at the 14 SNPs in Chinese_2025 cohort was performed using PLINK v1.9.0 [8]. Meta-analyses with East_Asian_2023 and European_2023 cohorts were performed using PLINK v1.9.0 [3]. All datasets were referenced to Genome Reference Consortium Human Build 37 (GRCh37). We performed an inverse variance weighted meta-analysis for the East Asian cohorts meta-analysis and the cross-ancestry meta-analysis. We also performed a random-effects meta-analysis for the latter; as expected, p-values were slightly larger with this method (results not shown).

### GWAS Power Calculations based on observational OR

We evaluated the power to detect associations between *C1GALT1* genetic variants known to influence serum Gd-IgA1 levels and IgAN, first in the previously published IgAN GWAS cohort [3] and secondly with the addition of the newly genotyped cohort. Power calculation was performed using the R package genpwr. The OR_SNP→IgAN_ required as an input for this was calculated as follows:

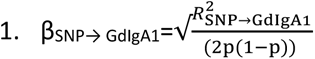

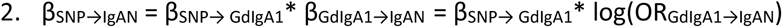

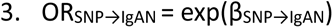

R^2^ is the previously derived variance in Gd-IgA1 explained by *C1GALT1* lead SNPs (range: 0.009–0.042). OR_GdIgA1→IgAN_ is the observational OR for IgAN per one standard deviation increase in standardised residuals of Gd-IgA1 after adjustment for age, sex, cohort, and total IgA levels (range: 1.49–1.56) as reported previously [4,5]. The alternate allele frequencies *p* for lead SNPs were derived from the dbSNP ALFA population frequencies for European and East Asian populations. We performed power calculations at a genome-wide level α=5x10^-8^ and a nominally significant α=0.01 to reflect testing of the prior hypothesis that elevated Gd-IgA1 levels cause IgAN.

### Mendelian Randomisation

We performed two-sample Mendelian randomisation (MR) analysis, with Gd-IgA1 level as the exposure variable and IgAN status as the outcome variable. Data for exposure cohorts are summarised in Table 1.

We analysed the data with two complementary MR methods: the Wald Ratio method using a single instrumental variable (IV) and inverse variance weighted approach (IVW) for the multiple IV MR. We sought IVs that were strongly associated with the exposure and likely to affect the outcome directly through the exposure. We subsequently tested for horizontal pleiotropy [9].

The Wald Ratio method was applied to the newly meta-analysed IgAN GWAS outcome data as our newly genotyped SNPs were all in the *C1GALT1* locus and somewhat correlated (r^2^>0.3). A lead exposure SNP (rs13226913) was identified in the European_2017 and British_2017 exposure cohorts (β=0.2512, p-value 8.27x10^-9^, F-statistic 16.1) and present in the European_2023 outcome data. An IVW-meta-analysed exposure estimate was used. This SNP is rarer and explains less variance in Gd-IgA1 levels in East Asian cohorts than European. The most significant SNP from the Chinese_2021 cohort was rs10238682 (β=-0.26, p-value 1.20×10^−9^. R^2^=0.037) but this was not present in the newly genotyped SNPs so we used SNP rs1008898 (β =-0.2282, p-value 9.39x10^-8^, F-statistic 31.0) which is highly correlated (r^2^=0.897) with it (Figure S1). We performed an IVW meta-analysis of these two MR estimates. Noting that this SNP was reasonably common and had a p-value of 3.53x10^-5^ for Gd-IgA1 in the British_2017 cohort (F-statistic 15.45), we estimated a cross-ancestry exposure effect and used this with the 3-way outcome effect estimate. Single IV MR was performed in R (see script appendix).

Orthogonally, the multiple IV approach was applied to the East_Asian_2023 IgAN outcome using the Chinese_2021 exposure data and the European_2023 IgAN outcome using the British_2017 exposure data [4]. To select valid instrumental variables, we identified SNPs present in both exposure and outcome datasets, excluded palindromic SNPs and eliminated highly correlated SNPs through linkage disequilibrium clumping and pruning with a clumping distance of 10kb and R^2^ <0.001, based on the 1000 Genomes Project [10]. We chose five SNPs in total with the smallest p-values for the exposure per chromosome above a genome-wide suggestive significance threshold of p<5×10^−5^. If more than one SNP was available on a chromosome the one with the highest F-statistic was chosen. To assess the strength of the selected instrumental variable we calculated the F-statistic with the following formula [11]:

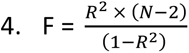

Rearranging [1.] above to calculate the R^2^, where N represents the sample size [11]. Sensitivity analyses included MR-Egger method and weighted median method, both of which allow and adjust for the potential effects of horizontal pleiotropy. The two multi-IV MR estimates were then meta-analysed. Calculations were performed using the TwoSampleMR package version 0.6.16.

MR power to detect an effect of Gd-IgA1 levels on IgAN risk was calculated using the observational OR_GdIgA1→IgAN_ estimate, variance explained in Gd-IgA1 levels by SNP (R^2^) and allele frequencies, using the formula derived by Burgess for a single IV with a binary outcome [12]. Calculations were done using R 4.3.0.

## Results

### GWAS Meta-analysis

We estimated the power to detect a genotype-phenotype association from 10,079 newly genotyped Chinese IgAN cases with 5,030 controls (Chinese_2025, Table 2). We also estimated power in the previously published GWAS (referred to here as Biethnic_2023, and its component cohorts European_2023 and East_Asian_2023) [3] and power from meta-analysis of these studies. We first calculated an expected OR_SNP→IgAN_ from the observational effect sizes of Gd-IgA1 level on IgAN for different cohort sizes and using the previously reported genetic variance explained in Gd-IgA1 (R^2^). These calculations were made using the previously reported, observed effect sizes of the association of Gd-IgA1 level on IgAN risk. Figure S2 shows that these power calculations are strongly affected by the resulting R^2^_SNP→IgAN_. For the biethnic group, the 2023 study had power of ∼0.5 to detect an association at a genome wide level, and was strongly powered to detect an association at more permissive levels (power for α=0.01 level, 0.998). The Chinese_2025 cohort on its own was modestly powered (0.43) to detect an association at α=0.01. Meta-analysis with East_Asian_2023 cohort has >90% power if testing a SNP with R^2^_SNP→IgAN_ = 3.7% as reported by Wang et al (2021) and much less if R^2^ = 0.9% as shown in Kiryluk (2017). The cross ethnic met-analysis is strongly powered even with the weaker SNP to detect an association at a genome-wide discovery threshold of α=5x10^-8^ (power=0.983).

**Table 2.** Power calculations for different cohorts, assuming the observational OR _GdIgA1→IgAN_ and previously noted variance explained in Gd-IgA1 to calculate an expected OR_SNP→IgAN._ a. Cohort sizes from Kiryluk et al 2023 [3]. b. R^2^ and observational OR _GdIgA1→IgAN_ from Kiryluk et al 2017 [5] (Supplementary Table 4 and Supplementary Figure 5) c. R^2^ and observational OR _GdIgA1→IgAN_ from lead SNP, Wang et al. 2021. d. Inferred from b. e. Calculated from the rs13226913 European and East Asian T allele frequencies in dbSNP, multiplied by proportions for mixed cohorts. f. Calculated from the rs10238682 East Asian T allele frequencies in dbSNP.

| Cohort | N (proportion of cases) | $R^2_{\text{SNP} \rightarrow \text{GdIgA1}}$ | Allele frequency | OR<sub>GdIgA1→IgAN</sub> (95% CI) | Expected OR<sub>SNP→IgAN</sub><sup>e</sup> | Power ($\alpha=0.01$) | Power ($\alpha=5 \times 10^{-8}$) | MR power ($\alpha=0.01$) | MR Power ($\alpha=5 \times 10^{-8}$) |
| --- | --- | --- | --- | --- | --- | --- | --- | --- | --- |
| Biethnic_2023 | 38897 (0.26)<sup>a</sup> | 0.022<sup>b</sup> | 0.418<sup>e</sup> | 1.53 (1.4 - 1.68)<sup>b</sup> | 1.095 | 0.998 | 0.506 | 0.998 | 0.502 |
| European_2023 | 26734 (0.21)<sup>a</sup> | 0.042<sup>b</sup> | 0.580<sup>e</sup> | 1.49 (1.31 - 1.72)<sup>b</sup> | 1.124 | 0.998 | 0.487 | 0.998 | 0.497 |
| East_Asian_2023 | 12163 (0.38)<sup>a</sup> | 0.009<sup>b</sup> | 0.062<sup>e</sup> | 1.56 (1.37 - 1.78)<sup>b</sup> | 1.131 | 0.382 | 0.001 | 0.375 | 0.001 |
| Chinese_2025 | 15109 (0.67) | 0.009<sup>b</sup> | 0.062<sup>e</sup> | 1.56 (1.37 - 1.78)<sup>b</sup> | 1.131 | 0.433 | 0.001 | 0.445 | 0.001 |
| East_Asian_2023, Chinese_2025 | 27272 (0.53) | 0.009<sup>b</sup> | 0.062<sup>e</sup> | 1.56 (NA)<sup>b</sup> | 1.131 | 0.811 | 0.023 | 0.815 | 0.024 |
| East_Asian_2023, Chinese_2025 | 27272 (0.53) | 0.037<sup>c</sup> | 0.480<sup>f</sup> | 1.53 (NA)<sup>c</sup> | 1.129 | 1.000 | 0.943 | 1.000 | 0.944 |
| East_Asian_2023, Chinese_2025, European_2023 | 54006 (0.37) | 0.025<sup>d</sup> | 0.319 | 1.53 (1.4 - 1.68)<sup>c</sup> | 1.107 | 1.000 | 0.983 | 1.000 | 0.983 |

Tests at the 14 SNPs across the *C1GALT1* locus in the new Chinese cohort showed no genome wide significant associations. Three were nominally significant at α=0.01 level (rs4263662(T), OR=1.083, p=0.0012; rs73045773(G), OR=0.894, p=0.0007; rs2190935(T), OR=1.093, p=0.0020). However, when comparing them to direction of effect of the allele on Gd-IgA1 levels, the effect for rs73045773(G) was discordant (i.e. the allele previously associated with increased Gd-IgA1 in the Chinese population was associated with *reduced* IgAN risk), and using European exposure data the other two were discordant (Table 3). Despite being adequately powered, no SNP reached genome wide significance in either meta-analysis. In the Chinese-only meta-analysis additionally SNP rs10259085(C) (OR = 1.057) was nominally significant at α= 0.01. In the cross-ancestry meta-analysis, two SNPs rs4263662(T) and rs2190935(T) were nominally significant at α=0.01, with OR=1.05 and 1.04 respectively. Notably, of these suggestively significant SNPs both show a discordant direction of effect on IgAN risk compared with the European exposure GWAS. Thus, of the four SNPs nominally significantly associated with IgAN in at least one of these analyses, only for rs10259085(C) was the effect concordant with the effect seen in both Gd-IgA1 GWASs.

**Table 3.** Association at 14 SNPs spanning *C1GALT1* in 15,109 Chinese subjects and meta-analysis with previous GWAS (Kiryluk et al 2023) [3]. Effect size of test allele on Gd-IgA1 level, where tested and p<0.05, in Chinese_2021 and British_2017 cohorts shown. IgAN association in European_2023 cohort not shown since p>0.01 for all. *Nominally significant at the level of p-value <0.01; **#** In strong linkage disequilibrium with lead SNP rs10238682 from Chinese_2021 cohort. **##** Discordant direction of allele effect compared with SNP→Gd-IgA1 in Chinese_2021 cohort **^^^** H1 Gd-IgA1 associated haplotype in British_2017 cohort. **^^^^** Discordant direction of allele effect compared with SNP→Gd-IgA1 in British_2017 cohort.

| Association tested |  |  | <u>Gd-IgA1 Association</u> |  | <u>IgAN Outcome Association</u> |  |  |  | Chinese_2025, East_Asian_2023, European_2023 |  |
| --- | --- | --- | --- | --- | --- | --- | --- | --- | --- | --- |
| Cohort |  |  | British_2017 | Chinese_2021 | Chinese_2025 |  | Chinese_2025, East Asian 2023 |  |  |  |
| SNP | Position | Test Allele | β (P) | β (P) | OR (95% CI) | P | OR (95% CI) | P | OR (95% CI) | P |
| rs4720724<sup>^</sup> | 7:7206760 | G | 0.2208 (4.6x10<sup>-7</sup>) | - | 0.9612 (0.8799 - 1.0501) | 0.3806 | 1.0208 (0.9571 - 1.0888) | 0.5311 | 1.0016 (0.9601 - 1.0449) | 0.9409 |
| rs758263<sup>^</sup> | 7:7221968 | T | 0.2543 (5.5x10<sup>-9</sup>) | 0.3001 (2.5x10<sup>-3</sup>) | 0.5938 (0.8686 - 1.0473) | 0.3215 | 1.0072 (0.9348 - 1.0853) | 0.8501 | 1.0054 (0.9572 - 1.0561) | 0.8295 |
| rs4263662<sup>^^</sup> # | 7:7227427 | T | -0.1431 (1.2x10<sup>-3</sup>) | 0.1953 (4.3x10<sup>-6</sup>) | 1.0831 (1.0320 - 1.1369) | 0.0012* | 1.0582 (1.0231 - 1.0946) | 0.0010* | 1.0467 (1.0184 - 1.0757) | 0.0011* |
| rs7788032 | 7:7243446 | G | - | - | 0.9017 (0.8088 - 1.0053) | 0.0621 | 0.9931 (0.9152 - 1.0777) | 0.8685 | 0.9868 (0.9403 - 1.0356) | 0.5892 |
| rs13226913<sup>^</sup> | 7:7246846 | T | 0.2512 (8.3x10<sup>-9</sup>) | 0.2499 (0.01) | 0.9633 (0.8767 - 1.0585) | 0.4368 | 1.0219 (0.9528 - 1.0961) | 0.5436 | 1.0076 (0.9649 - 1.0522) | 0.7309 |
| rs2108790 | 7:7255479 | C | - | - | 0.9332 (0.8375 - 1.0398) | 0.2100 | 1.0053 (0.9276 - 1.0895) | 0.8972 | 0.9900 (0.9437 - 1.0384) | 0.6788 |
| rs10259085<sup>^</sup> | 7:7268431 | C | 0.1558 (4.2x10<sup>-4</sup>) | 0.1596 (5.7x10<sup>-4</sup>) | 1.0630 (1.0091 - 1.1199) | 0.0215 | 1.0565 (1.0186 - 1.0959) | 0.0032* | 1.0344 (1.0059 - 1.0637) | 0.0177 |
| rs9639031 | 7:7273155 | T | - | 0.1575 (7.0 x 10<sup>-4</sup>) | 1.0533 (0.9997 - 1.1099) | 0.0515 | 1.0472 (1.0095 - 1.08630) | 0.0137 | 1.0320 (1.0034 - 1.0614) | 0.0278 |
| rs73045773<sup>##</sup> | 7:7273424 | G | - | 0.1475 (6.4x10<sup>-3</sup>) | 0.8940 (0.8376 - 0.9542) | 0.0007* | 0.9423 (0.9015 - 0.9850) | 0.0086* | 0.9597 (0.9252 - 0.9955) | 0.0279 |
| rs1008898<sup>^</sup> # | 7:7273559 | G | 0.1821 (3.5x10<sup>-5</sup>) | 0.2282 (9.4x10<sup>-8</sup>) | 1.0471 (0.9982 - 1.0984) | 0.0592 | 1.0369 (1.0028 - 1.0721) | 0.0339 | 1.0274 (1.0002 - 1.0553) | 0.0483 |
| rs2190935<sup>^^</sup> | 7:7283017 | T | -0.1078 (1.5x10<sup>-2</sup>) | - | 1.0930 (1.0330 - 1.1566) | 0.0020* | 1.0602 (1.0194 - 1.1027) | 0.0035* | 1.0425 (1.0122 - 1.0736) | 0.0056* |
| rs1047763 | 7:7283569 | G | - | - | 1.0206 (0.9732 - 1.0704) | 0.4005 | 1.0310 (0.9968 - 1.0663) | 0.0758 | 1.0260 (0.9983 - 1.0546) | 0.0662 |
| rs5010183 | 7:7286198 | C | - | - | 0.9323 (0.8415 - 1.0330) | 0.1804 | 1.0033 (0.9304 - 1.0820) | 0.9317 | 1.0012 (0.9574 - 1.0470) | 0.9581 |
| rs35463640 | 7:7291646 | A |  | 0.2756 (5.2x10<sup>-3</sup>) | 0.9438 (0.8518 - 1.0457) | 0.2685 | - | - | - | - |

### Mendelian Randomisation

For given sample sizes, two-sample MR may allow greater power to detect an association between an exposure and an outcome than the power to detect an association directly between a SNP and an outcome, dependent on the variance of exposure explained by the SNP. The framework also allows a direct estimate of the exposure-outcome effect (i.e. OR_GdIgA1→IgAN)_). Using the observational OR_GdIgA1→IgAN_ estimate, we calculated that MR would be strongly powered to detect an association in the each of the cohorts (Table 2).

We performed a two-sample Wald ratio MR study to estimate the likely size of any causal effect of Gd-IgA1 levels on IgAN risk. We use the strongest available instrument for each population and then meta-analysed these instruments; and also show the result if a combined effect of one instrument was used for both populations, with the cross-ancestry meta-analysis (Table 4).

**Table 4.** Wald Ratio Method Mendelian Randomisation study with rs13226913 and rs1008898 as a single genetic instrumental variable.

| SNP | Position /<br>Effect allele | Exposure cohort | Outcome cohort | N of outcome cohort<br>(proportion of cases) | Estimated exposure<br>$R^2$ SNP → GdIgA1 | $\beta_{\text{GdIgA1} \rightarrow \text{IgAN}}$ (SE) | $\text{OR}_{\text{GdIgA1} \rightarrow \text{IgAN}}$ per 1 SD Gd-IgA1 (95%CI) | P |
| --- | --- | --- | --- | --- | --- | --- | --- | --- |
| rs13226913 | 7:7246846 / T | British_2017,<br>European_2017 | European_2023 | 26734 (0.21) | 0.0320 | -0.0961 (0.1209) | 0.9083 (0.7175 - 1.1500) | 0.4264 |
| rs1008898 | 7:7273559 / T | Chinese_2021 | Chinese_2025,<br>East_Asian_2023 | 27272 (0.54) | 0.0260 | 0.1586 (0.0805) | 1.1719 (1.0001- 1.3721) | 0.0487 |
| Meta-analysis of these<br>estimates |  |  |  | 54006 (0.37) |  | 0.0804 (0.0670) | 1.0837 (0.9504 - 1.2357) | 0.2302 |
| rs1008898 | 7:7273559 / T | Chinese_2021,<br>British_2017 | Chinese_2025,<br>East_Asian_2023,<br>European_2023 | 54006 (0.37) | 0.0202 | 0.1313 (0.0693) | 1.1403 (0.9955 -1.3061) | 0.0581 |

The Chinese meta-analysis showed an OR of 1.17 with a wide 95% CI (1.00-1.37), with a p-value of 0.049. The European and cross-ancestry meta-analyses showed smaller or reverse effects of the Gd-IgA1 on IgAN risk with wider confidence intervals.

As an orthogonal MR method to leverage more information from the exposure datasets, we performed multi-variable MR on European and East Asian populations and then meta-analysed the effect estimate. We selected five independent SNPs (only one of which was in the *C1GALT1* locus) with the most significant p-value from our summary statistics of GWAS of Gd-IgA1 levels for both populations and performed a multiple IV MR within the Kiryluk 2023 dataset with the corresponding ethnicities (Table 5 and Figure 2, Table S2). MR-IVW analysis did not demonstrate any evidence of association of Gd-IgA1 level with the risk of IgAN. Further sensitivity analyses with MR-Egger and Weighted median methods which allow violation of some MR assumptions, did not show a significant association of genetically predicted Gd-IgA1 level with the risk of IgAN (Table 5).

**Figure 2.**
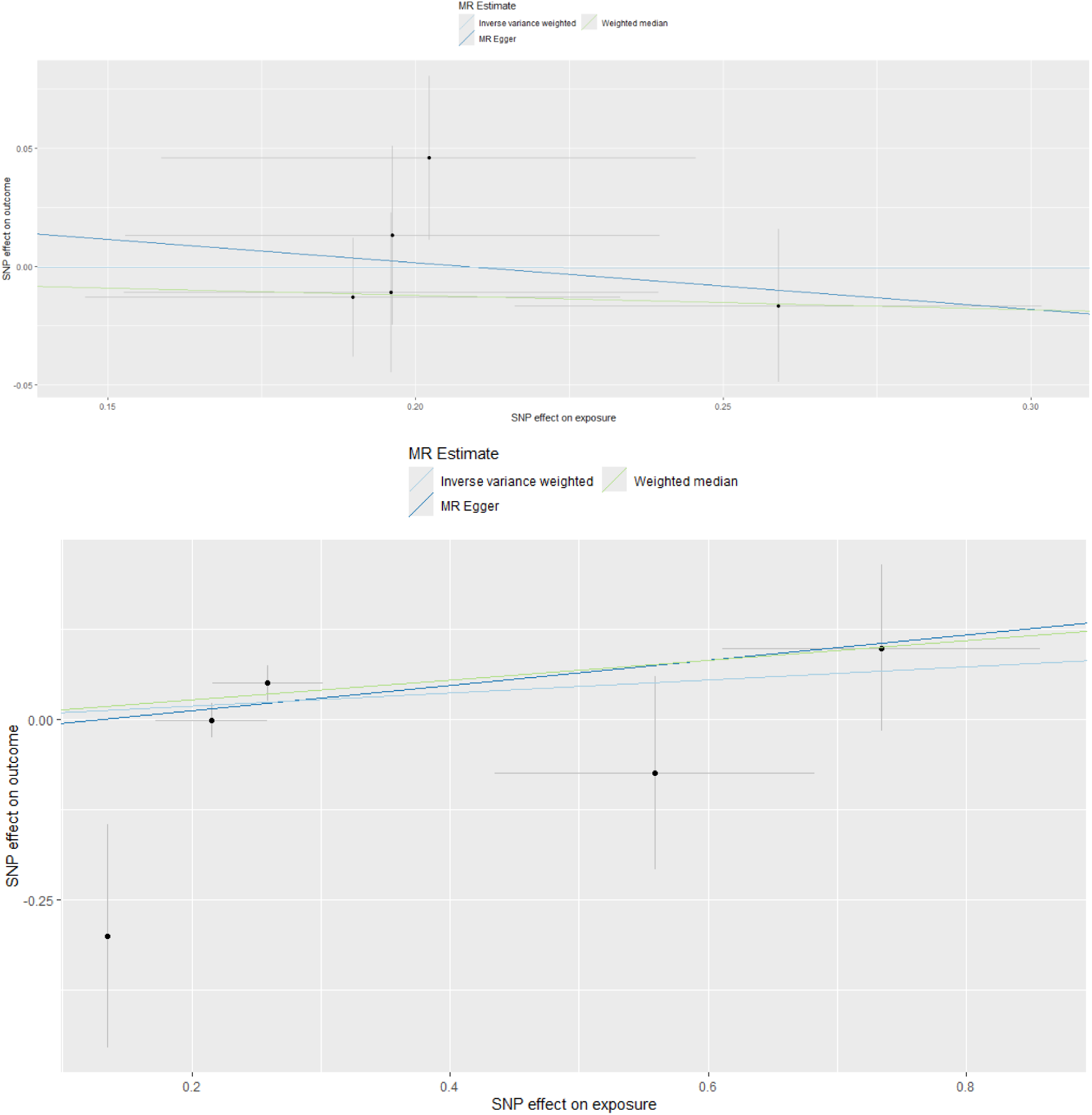
Scatter-plots showing the effect estimate of each instrumental variable on Gd-IgA1 (exposure) and IgAN risk (outcome) in the multiple IV MR (Top: European; Bottom: East Asian).

**Table 5.** Multiple instrumental variable Mendelian Randomisation. Five uncorrelated SNPs were selected from each of the exposure cohort according to their strength of association to Gd-IgA1 level. British_2017: rs1008897 (p=2.83x10^-9^), rs1344616 (p=4.01x10^-6^), rs519380 (p=7.70x10^-6^), rs294657 (p=7.86x10^-6^), rs12140760 (p=1.53x10^-5^). Chinese_2021: rs10238682 (p=1.20 x10^-9^), rs7856182 (p=2.38 x10^-9^), rs3782690 (p=7.73 x10^-7^), rs142263068 (p=6.37 x10^-6^), rs9513048 (p=3.91 x10^-7^). IVW, MR-inverse variance weighting method; Egger, MR-Egger method; Weighted median method

| Exposure Cohort | Outcome Cohort | MR-analysis | $\beta_{\text{GdIgA1} \rightarrow \text{IgAN}}$ (SE) | $\text{OR}_{\text{GdIgA1} \rightarrow \text{IgAN}}$ per 1 SD Gd-IgA1 (95%CI) | P |
| --- | --- | --- | --- | --- | --- |
| British_2017 | European_2023 | IVW | -0.0019 (0.0681) | 0.9981 (0.8733 – 1.1408) | 0.9783 |
|  |  | Egger | -0.1977 (0.5567) | 0.8206 (0.2756 – 2.4433) | 0.7460 |
|  |  | Weighted median | -0.0612 (0.0828) | 0.9407 (0.7998 - 1.1064) | 0.4599 |
| Chinese_2021 | East_Asian_2023 | IVW | 0.0975 (0.0629) | 1.1024 (0.9744 – 1.2471) | 0.1215 |
|  |  | Egger | 0.1157 (0.2391) | 1.1226 (0.7026 – 1.7938) | 0.6764 |
|  |  | Weighted median | 0.1369 (0.0777) | 1.1467 (0.9846 - 1.3354) | 0.0783 |
| IVW Meta-analysed of these estimates |  |  | 0.0621 (0.0487) | 1.0641 (0.9672 – 1.1701) | 0.2024 |

## Discussion

The “four-hit hypothesis” offers a testable model incorporating what is currently know about the pathogenesis of the disease. Despite observational data showing an association between Gd-IgA levels and IgAN risk, at least seven large-scale GWAS for IgAN to identify susceptibility loci, including the largest GWAS recently published, did not demonstrate association at *C1GALT1*, the main genetic determinant of Gd-IgA1 levels, at genome-wide significance threshold [3, 13–18]. Our regional meta-analysis with 20,236 cases (10,146 of which are new), did not detect evidence of genetic effects on IgAN risk at the *C1GALT1* locus at a genome-wide significance level, despite being adequately powered to detect an effect of a magnitude consistent with the reported effect of Gd-IgA1 on IgAN risk from observational studies.

In a Chinese-only meta-analysis (case n=14,469), four SNPs showed evidence for a small effect on IgAN risk at a nominal α=0.01 level. The historic scientific literature contains numerous candidate gene association studies with borderline statistically significant evidence of association that have not withstood replication analyses, indicating the conservative statistical approach to interpreting such studies needed to avoid Type 1 errors [19,20]. In this case, the inclusion of the European cohort attenuated these signals, contrasting with the observed association of Gd-IgA1 levels with IgAN that is consistent across all cohorts studied. Moreover, some of these signals were inconsistent with expected direction of effect: alleles associated with increased Gd-IgA1 levels in at least one of the populations were associated with decreased IgAN risk. This is inconsistent with the hypothesis that genetic risk factors for elevated Gd-IgA1 are risk factors for IgAN.

We performed two types of Mendelian Randomisation analysis to quantify the evidence for a causal role of Gd-IgA1 in IgAN. The MR approach also allows estimation of the magnitude of any effect of Gd-IgA1 on IgAN risk. The single IV MR on the largest Chinese cohort is consistent with a causal effect, but this is not seen in the European cohort and does not survive meta-analysis. This association was not replicated in in the multiple IV MR analyses we performed. Importantly, in this analysis, the effect size estimates are lower than the observational effect estimates, and the upper bounds of the ORs were 1.37 and 1.24 by the two methods, respectively, just overlapping the lower bounds of effect estimate from observational data. Overall, our MR analyses showed that the observed strength of the association cannot be explained by a causal relationship between Gd-IgA1 levels and IgAN.

In both association analyses and MR frameworks our study was well-powered to detect associations consistent with the observed effect size of Gd-IgA1 levels on IgAN risk. We do not find evidence for a causal effect of this magnitude. We cannot exclude that the magnitude of this observed effect size is overstated and that a smaller effect could be detected by larger IgA GWAS sample size. However, this is unlikely to find consistent evidence across cohorts of genetic effects on IgAN at the *C1GALT1* locus, given the observed discordance of effects that we have demonstrated.

These genetic findings are supported by several other findings that argue against Gd-IgA1 playing a causal role in the disease. Previous studies have shown that Gd-IgA1 level were elevated in some asymptomatic first-degree relatives of patients with IgAN [21] and its circulating level did not correlate with disease activity or progression, despite being clearly elevated in patients with IgAN [22]. Furthermore, the discrepancies of Gd-IgA1 level between different ethnicities, especially taking into account the prevalence of IgAN across the globe, shows that elevated Gd-IgA1 levels cannot directly explain the higher prevalence of IgAN in East Asia [4]. Plausible alternative hypotheses to explain the observed association of Gd-IgA1 levels with IgAN risk include that the observed association is a consequence of IgAN (reverse causation), or that Gd-IgA1 is elevated by other factors (such as mucosal inflammation) that also increase risk of IgAN (i.e. confounding).

Our study has some strengths. First, the additional of our large cohort to the current largest published IgAN GWAS has significantly augmented the power to detection an association. From the power calculation, our cohort clearly has sufficient power to detect an association, at a robust α of 5x10^-8^. We have tested the association in two populations with population-appropriate instrumental variables and meta-analysed these results. Second, our genetic instruments have F-statistics above 10, which reduce the risks of weak instrumental bias. MR methods are often criticised for evoking false positive results by not accounting adequately for confounding by pleiotropy or reverse causation. Importantly, however, the key inference of our study differs from the typical MR claim of *causal association*: here, we demonstrate robust *lack of association* in several adequately powered analyses.

Our study has several limitations. First, we selectively genotyped only the *C1GALT1* locus rather than performing whole genome sequencing or genotyping; we increased power to detect IgA associations only at this locus and may miss associations at other loci relevant to both Gd-IgA1 and IgAN. However, *C1GALT1* is the strongest known genetic determinant of Gd-IgA1; if no association with IgAN is seen here it is unlikely to be found elsewhere. Indeed, in the light of our findings, if an association with IgAN were found at other loci that are weaker determinants of Gd-IgA1, such as *GALNT12* [7] or *C1GALT1C1*, this would, by definition, represent an example of horizontal pleiotropy – i.e. the association with IgAN would be through pathways other than Gd-IgA1 levels. Finally, our MR framework assumed a linear relationship between Gd-IgA1 and IgAN risk. It is challenging to model power to detect gene-gene interactions (as proposed by Wang et al [7]) or gene-environment interactions; however the absence of evidence from the GWAS meta-analysis argues against the presence of strong enough such effects to explain the phenotypic association of Gd-IgA1 with IgAN.

In conclusion, despite seeking (using three genetic approaches) evidence of a causal effect of elevation of Gd-IgA1 on IgAN, our genetic data do not support the hypothesis that the well-observed phenotypic association between Gd-IgA1 and IgAN is attributable to causation. While it remains possible that genetic evidence from larger studies will uncover evidence consistent with a smaller causal effect it is reasonable to propose the hypothesis that Gd-IgA1 elevation represents an epiphenomenon rather than a causal driver of the disease. This hypothesis is testable, for example by using an interventional study design as reported by Wu et al (manuscript submitted).

Resolution of this issue has potentially important implications for therapeutic development: strategies specifically using Gd-IgA1 as the sole biomarker of impact on IgAN pathogenesis may over or underestimate their true disease modifying potential. Furthermore, ranking treatments based on the achieved magnitude of reduction in Gd-IgA1 could result in misleading conclusions on the disease modifying capabilities of new treatments in IgAN.

## Data Availability

All the data are presented in full in the manuscript and the summary statistics are downloaded from their respective papers (See data availability links).

https://doi.org/10.1038/s41588-023-01422-x

https://doi.org/10.1681/ASN.2016091043

https://doi.org/10.1371/journal.pgen.1006609

https://doi.org/10.1681/ASN.2020060823

## Acknowledgements and funding

GTD is supported by a Kidney Research UK Fellowship. DPG is supported by the St Peter’s Trust for Kidney, Bladder and Prostate Research.

## Declaration

The authors declare no conflict of interest.

## Data Sharing Statement

All the data are presented in full in the manuscript and the summary statistics are downloaded from their respective papers [3–5, 7]. Only publicly available open-source software was used in the analyses; there was no custom software.

## Author Contributions

DPG and JB conceived the study; WWSF and GTD performed the analyses with assistance from OS-A and X-JZ; WWSF, GTD and DPG wrote the manuscript which was revised and approved by all the authors.

## Supplementary files

**Figure S1.**
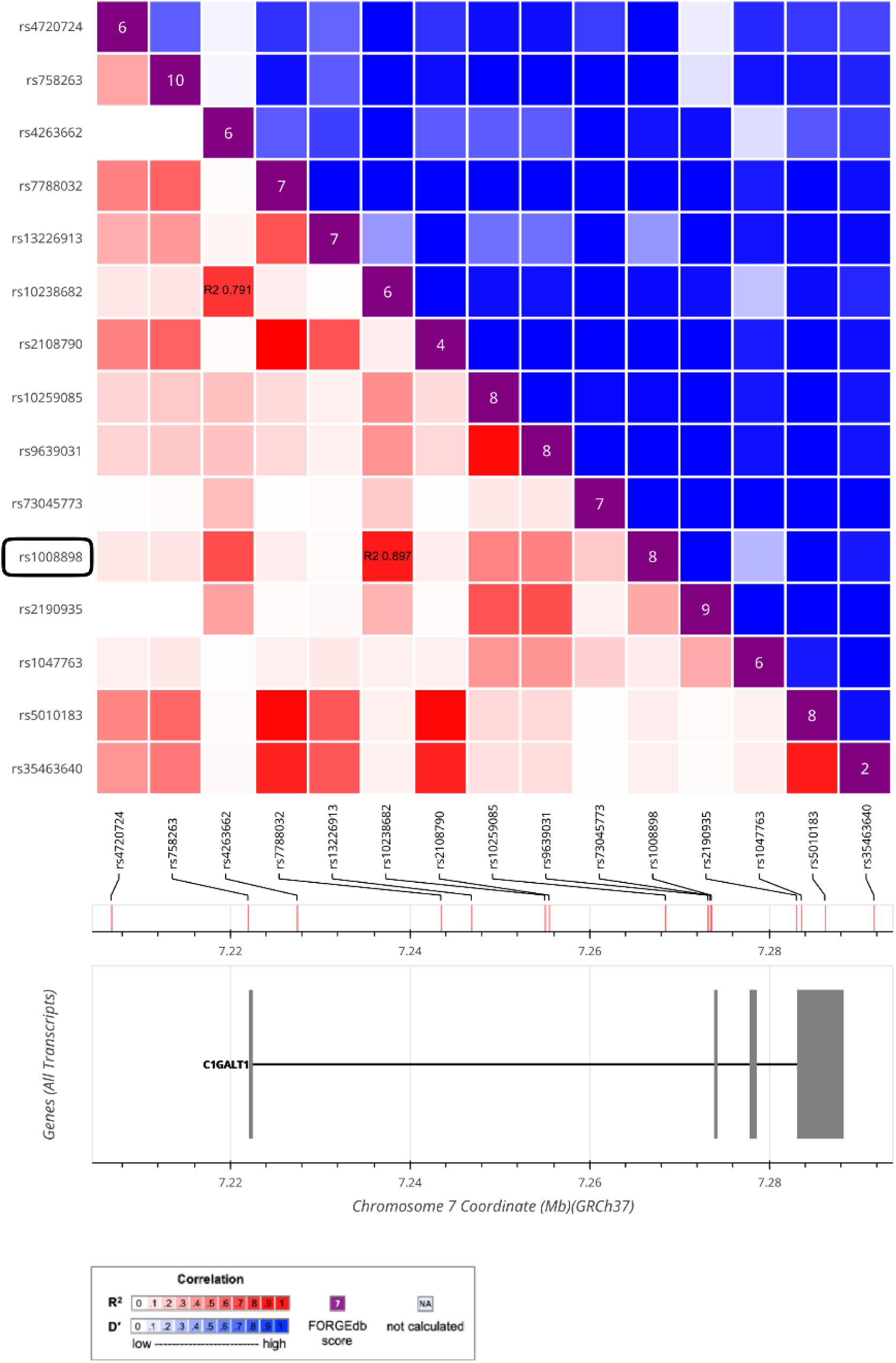
LD matrix of *C1GALT1* showing that rs10238682 is highly correlated with rs4263662 (r^2^ = 0.791) and rs1008898 (r^2^ = 0.897) in Chinese population.

**Figure S2.**
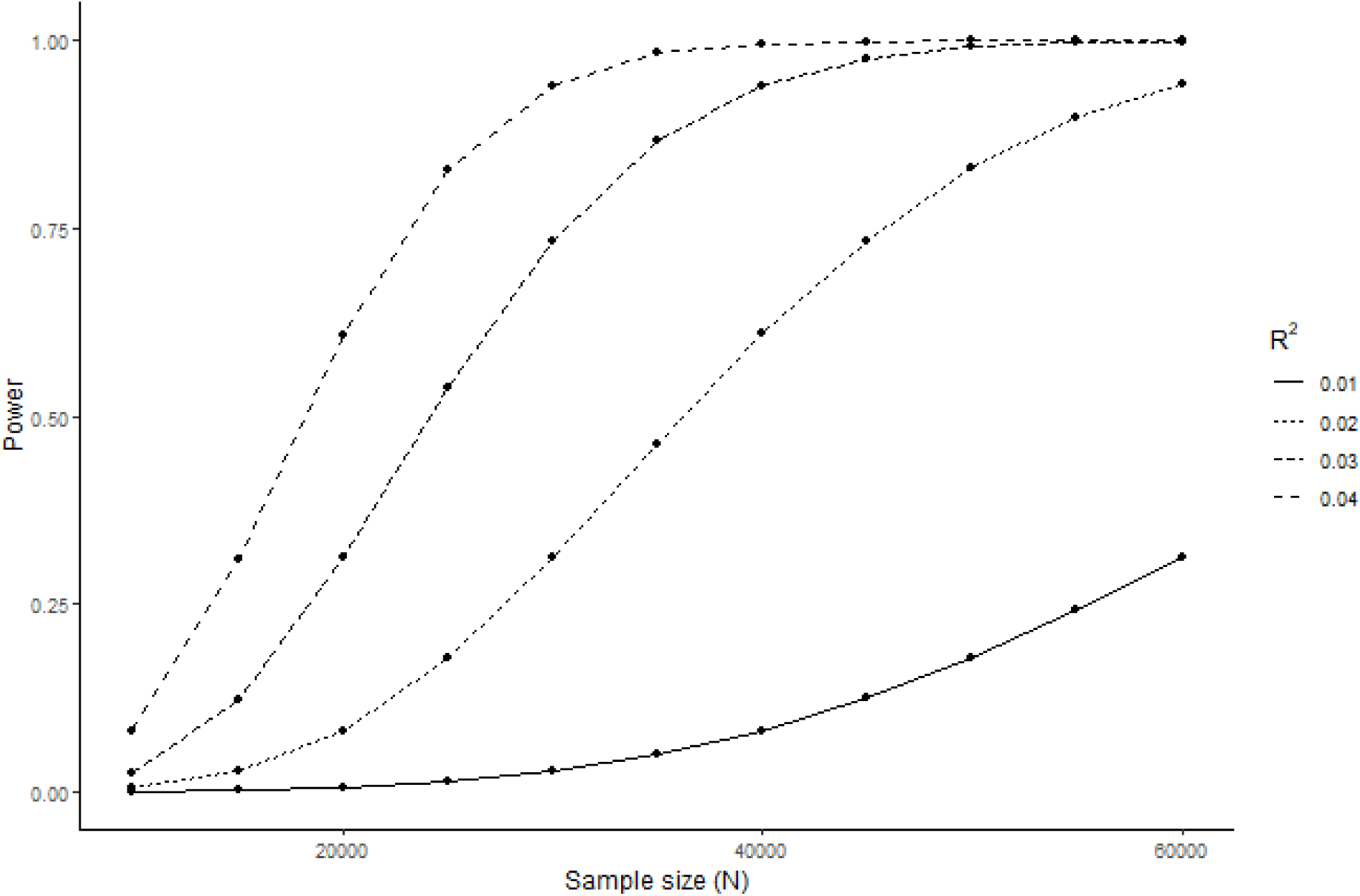
Power curve for logistic regression of SNP→outcome, under an additive assumption, based on a fixed exposure→outcome OR = 1.5 with differing SNP→exposure variance R^2^.

**Table S1.**
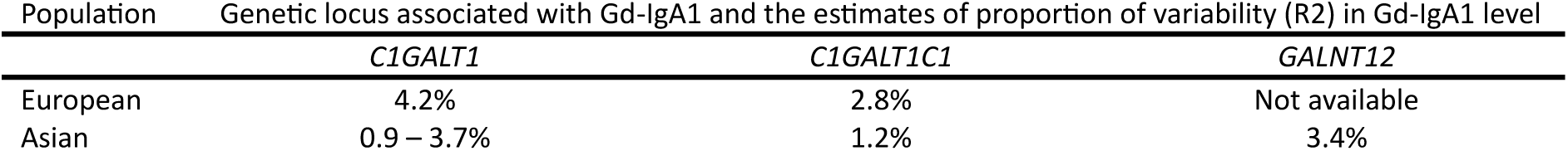
Estimates of proportion of variability (R2) in Gd-IgA1 level explained by genome-wide significant loci associated with Gd-IgA1 [4,5,7].

**Table S2.** Five independent and uncorrelated SNPs used for the multiple instrumental variable Mendelian Randomisation.

| SNP | Chromosome | Position / Effect allele | $\beta_{\text{SNP} \rightarrow \text{GdIgA1}}$ | p-value |
| --- | --- | --- | --- | --- |
| British_2017 |  |  |  |  |
| rs1008897 | 7 | 7273440 / G | 0.2590 | $2.83 \times 10^{-9}$ |
| rs1344616 | 8 | 69971497 / G | -0.2022 | $4.01 \times 10^{-6}$ |
| rs519380 | 11 | 66409409 / A | -0.1963 | $7.70 \times 10^{-6}$ |
| rs294657 | 2 | 123610641 / A | 0.1961 | $7.86 \times 10^{-6}$ |
| rs12140760 | 1 | 173429995 / A | -0.1899 | $1.53 \times 10^{-5}$ |
| Chinese_2021 |  |  |  |  |
| rs10238682 | 7 | 7255017 / G | -0.2588 | $1.20 \times 10^{-9}$ |
| rs7856182 | 9 | 101633312 / T | 0.7342 | $2.38 \times 10^{-9}$ |
| rs3782690 | 12 | 106464063 / G | 0.2153 | $7.73 \times 10^{-7}$ |
| rs142263068 | 5 | 94067448 / C | 0.5587 | $6.37 \times 10^{-6}$ |
| rs9513048 | 13 | 19989930 / G | 0.6911 | $3.91 \times 10^{-7}$ |

## R Script appendix

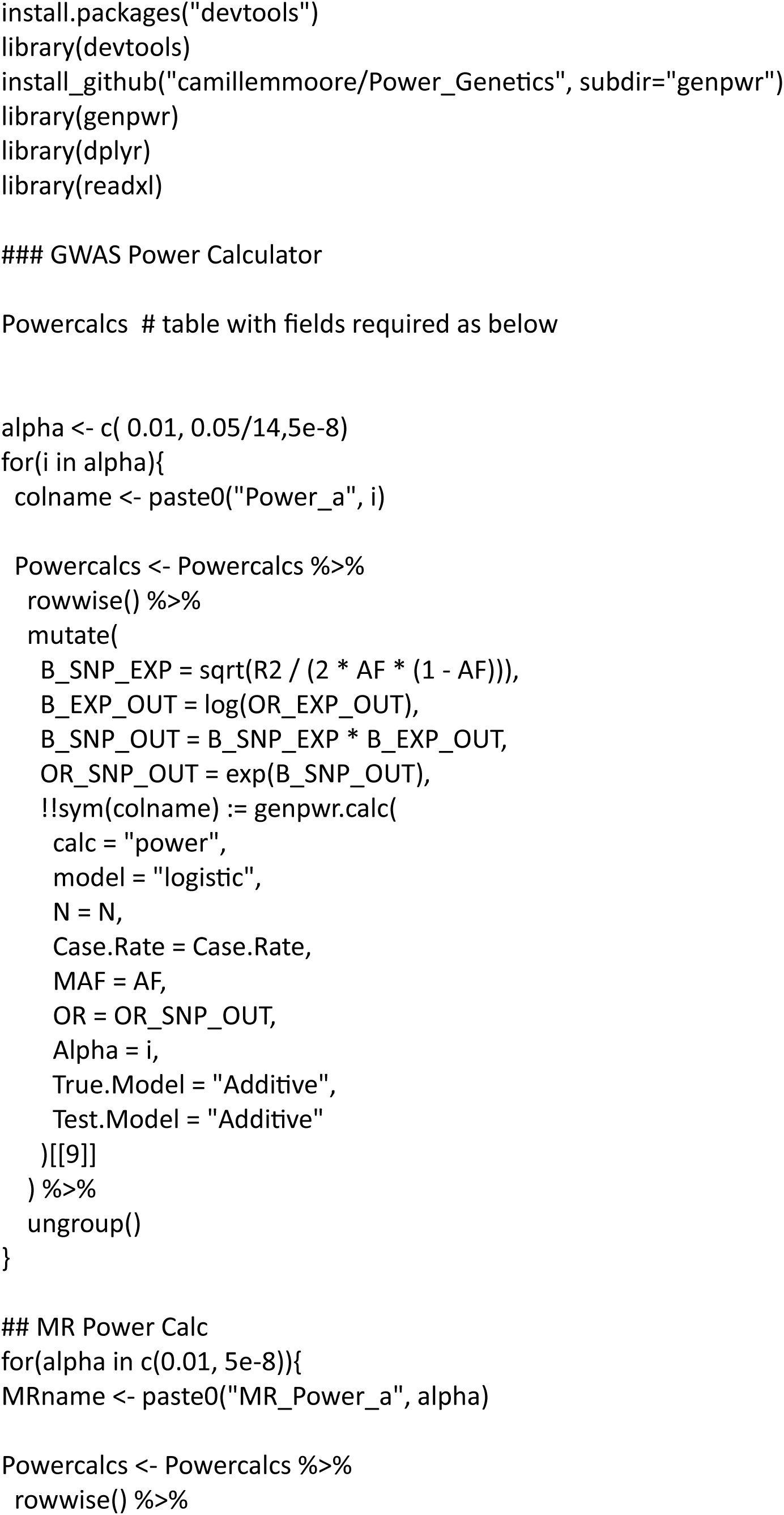
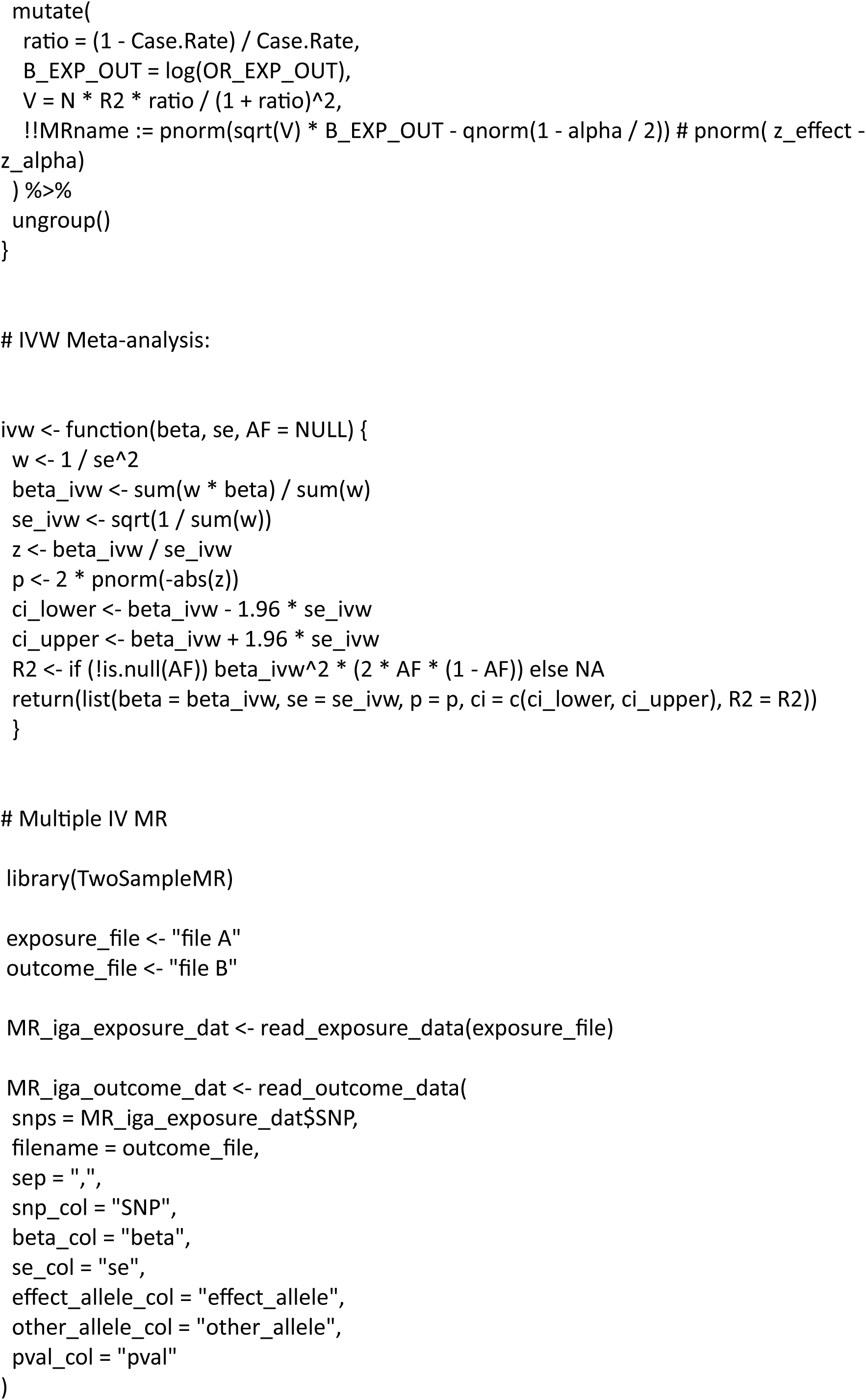
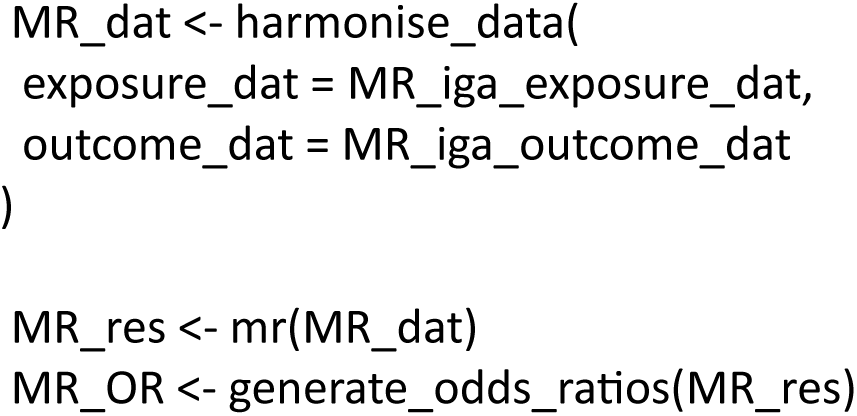

